# The United States CADASIL Consortium: Baseline Findings from a Natural History Study

**DOI:** 10.64898/2026.09.20.26363512

**Authors:** Jane S. Paulsen, H. Jeremy Bockholt, William H. Adams, Deven K. Burks, Jennifer J. Majersik, Stephen P. Salloway, Songmi Lee, Myriam Fornage, Cara Joyce, Jamie L. Elliott, Edward D. Huey, Megan R. Caruso, Suman Jayadev, José Biller, Helmi L. Lutsep, Sara J. Doyle, David S. Liebeskind, Kelsey Eklund, Imama A. Naqvi, Arash Salardini, Sudha Seshadri, Jason D. Hinman, Lisa C. Krishnamurthy, Barbara L. Fischer, Michael D. Geschwind, the United States CADASIL Consortiusm

## Abstract

**Background and Objectives:** Vascular contributions to cognitive impairment and dementia (VCID) represent the second leading cause of dementia and a common comorbidity for reduced functional capacity in many individuals, but clinical management and clinical trial readiness are severely hindered by extreme phenotypic and mechanistic heterogeneity. This study establishes the baseline clinical, functional, and multimodal biomarker characteristics of the first United States (US) cohort of Cerebral Autosomal Dominant Arteriopathy with Subcortical Infarcts and Leukoencephalopathy (CADASIL), serving as a single-cause, monogenic small vessel disease model for sporadic VCID.

**Methods:** Cross-sectional analysis of baseline data collected from October 2022 through January 2025 from a longitudinal cohort at 12 US enrollment sites. Adults from families with a documented *NOTCH3* variant were recruited through clinical referrals, medical record review, advocacy organizations, and community outreach. Exclusion criteria included other conditions that prevented interpretation of findings and modified Rankin Scale (mRS) scores greater than 3. Central genetic characterization assigned persons with a pathogenic or likely pathogenic *NOTCH3* variant to low-, medium-, or high-risk tiers based on epidermal growth factor-like repeat domains. Standardized assessments included clinical histories, exams, objective cognitive and physical assessments, self-reported, companion-reported, and clinician-rated measures, MRI, genetics, and proteomics acquired under harmonized protocols. Group comparisons used Fishers exact and Kruskal–Wallis tests. Multivariable regression used adjustment for age, sex, race, and education.

**Results:** Of 560 participants completing baseline visits, 46 had variants of unknown significance, and 55 had pending genetic analyses. The analytic cohort included 343 persons with a CADASIL-causing *NOTCH3* variant and 116 non-carrier family controls. Median age was 47.9 years (IQR 38.7–59.1), median education was 16 years (IQR 14– 17), 62.5% were female, and 91.4% were White. Carriers were classified in CADASIL risk tiers: low (7.0%), medium (18.4%), or high (74.6%). Compared with controls, adjusted odds of classification into a more impaired CDR category were higher in all tiers (OR 2.55-5.08, 95% CI 1.22–11.34).

**Discussion:** The US CADASIL Consortium has established a large, genetics-confirmed cohort spanning presymptomatic to moderate disease stages, providing a robust, harmonized platform to define early biomarker signatures and endpoints for targeted VCID therapies.

## Introduction

Vascular contributions to cognitive impairment and dementia (VCID) are the second leading cause of dementia globally and the most frequent co-occurrence in Alzheimer disease-related dementias (ADRD).^1,2^ Beyond aging, vascular-driven cognitive decline manifests in many conditions, including diabetes, multiple sclerosis, hypo- and hypertension, and systemic infections. Despite massive public health burden, clinical management of VCID and development of disease-modifying therapies are limited by extreme mechanistic heterogeneity and clinical variation among affected individuals. Insights derived from historical cohorts (e.g., Framingham Study, MarkVCID) provide critical evidence,^3,4^ but early pathogenic pathways remain confounded by mixed etiologies and variable risk profiles.

To isolate the mechanisms of VCID and establish clinical trial readiness, monogenic small vessel disease (SVD) models offer a powerful clinical window. Cerebral Autosomal Dominant Arteriopathy with Subcortical Infarcts and Leukoencephalopathy (CADASIL) is an inherited SVD caused by pathogenic variants in the *NOTCH3* gene. Characterized by recurrent subcortical strokes, sensory/motor deficits, psychiatric disturbances, and progressive cognitive decline, CADASIL serves as an effective model for VCID.^5^ Critically, because individuals with a CADASIL-causing *NOTCH3* gene variant can be identified and evaluated decades before overt clinical symptoms begin, this population helps researchers systematically map the earliest neurovascular and biofluid alterations driving this SVD.^5,6^

Current reviews of CADASIL indicate that more than 280 *NOTCH3* variants have been documented, including the original cysteine-altering missense mutations as well as splice-site mutations, small deletions/insertions, duplications, homozygous variants, loss-of-function mutations, and cysteine-sparing variants.^7^ Thus, diverse genotypes and the broad heterogeneity of the resulting phenotypes have challenged conventional diagnostic criteria for CADASIL. Researchers worldwide have responded to this conundrum with various efforts to bin the observed variants into lumped “risk status” as well as a recent call to designate the disease as a “spectrum”.^8,9^ Though most data come from cross-sectional hospital cohorts, stroke clinics, genetic databases, and research groups, over 20 countries have established or are currently developing CADASIL registries to advance knowledge of VCID heterogeneity contributing to clinical care challenges.^10^

Most notably, *NOTCH3* variants responsible for CADASIL are far more prevalent than historically recognized, appearing in up to 1 in 450 individuals in global population biobanks and frequently presenting with milder phenotypes.^11,12^ International studies indicate that CADASIL accounts for a substantial proportion of general VCID population—exceeding 1% in regions such as Taiwan.^13^ Because CADASIL is historically considered a rare disorder, patients with milder symptoms or atypical family histories are routinely missed. Careful characterization of pathogenic *NOTCH3* variants in a United States cohort may be helpful to lower diagnostic barriers, improve clinical recognition, and clarify pathologies in mixed dementia.

Furthermore, extensive white matter hyperintensities (WMH) triple the risk of future stroke and double the risk of developing dementia.^14^ Despite these stark risks, because WMH are ubiquitous in aging populations and linked to diverse etiologies (e.g., hypertension, lifestyle factors, multiple sclerosis, late-life depression), they are frequently dismissed in routine neurological practice as “incidental” or “silent”. Utilizing a monogenic SVD model across the life course isolates the natural history of WMH accumulation, equipping clinicians with actionable parameters to recognize when subclinical white matter changes demand aggressive diagnostic or therapeutic intervention.

Elucidation of genotype–phenotype correlations and their mechanisms is essential to explain the clinical heterogeneity of CADASIL. The exact mechanism of the disorder resists explanation despite the identification of the causative gene more than 25 years ago. This suggests that *NOTCH3* pathogenic variants cause more than just classical cerebral SVD.

To address these knowledge gaps, we established the United States CADASIL Consortium (USCC), the first unified, multi-site US cohort with a known monogenic cause of VCID.^6^ This article describes the USCC’s infrastructure, genotypic representation, baseline clinical profiles, and multimodal biomarker acquisition. By deeply phenotyping a large cohort spanning presymptomatic to moderate disease stages, these baseline findings lay another stone in the groundwork for clinical trial readiness, early diagnosis, and targeted therapies for both monogenic and sporadic VCID.

## Methods

### Study Design and Setting

This cross-sectional analysis uses baseline data from the USCC, an ongoing, longitudinal natural history study tracking individuals at baseline, 18 months, and 36 months. Travel was covered by the grant, and participants were recruited across the country to one of 12 enrollment sites. These data were collected between October 2022 and January 2025. This study followed the Strengthening the Reporting of Observational Studies in Epidemiology (STROBE; **Figure 1**) reporting guideline for cross-sectional studies.^15^

**Figure 1.**
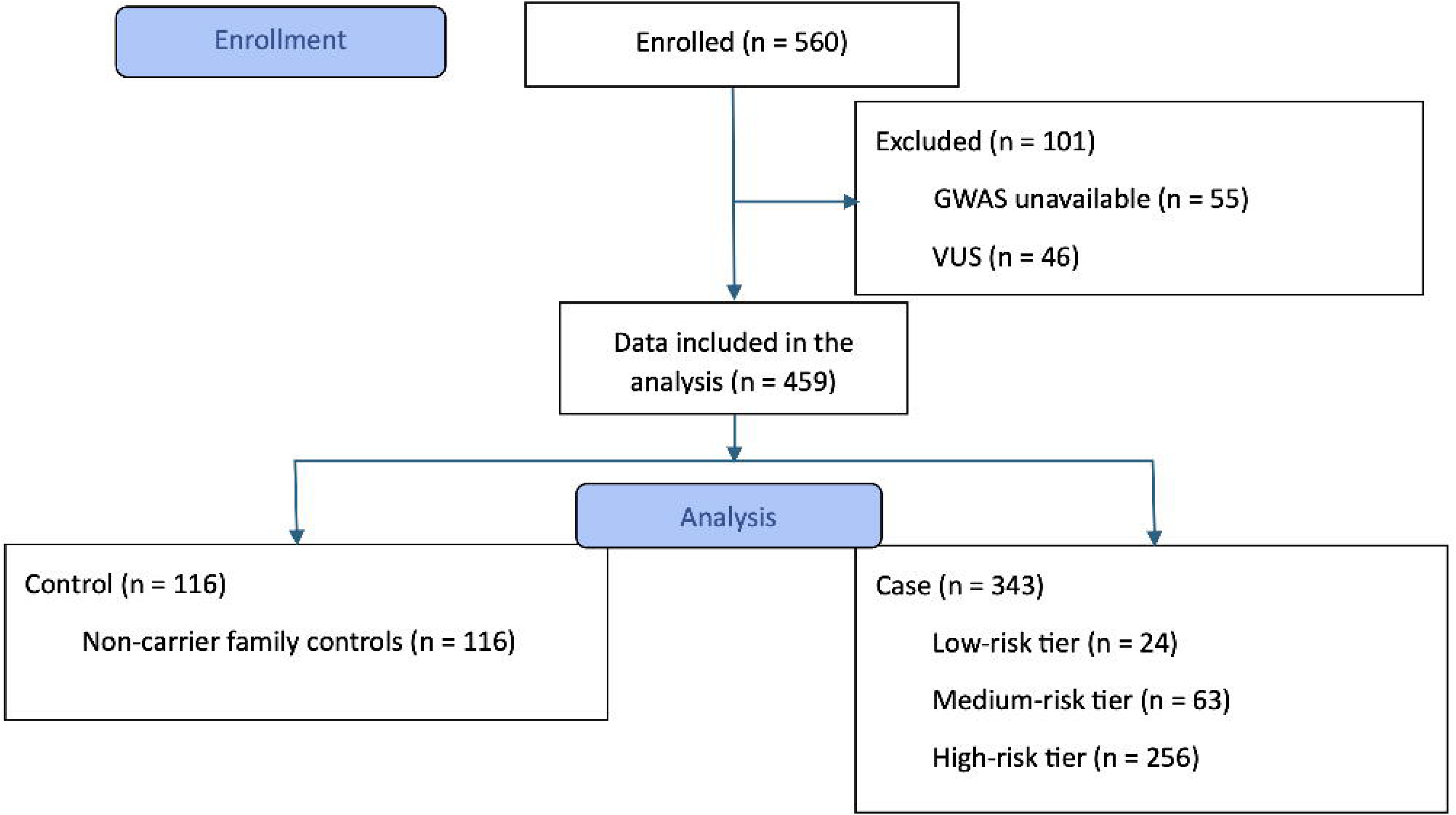
STROBE flow diagram for USCC cohort.

### Standard Protocol Approvals and Consents

The study protocol, recruitment materials, and consent forms were approved by the Western Clinical Group (WCG), a centralized institutional review board. All participants provided written informed consent before any study-related procedures. The study is registered at ClinicalTrials.gov (NCT05677880). Interested persons were allowed to enroll without individual prior genetic documentation if they possessed a documented or suspected pathogenic *NOTCH3* variant in any family member based on clinical and/or neuroimaging phenotype. All USCC participants were offered genetic counseling: 32% accepted this option, and a small proportion of these remained blinded to their genetic status, with visits conducted in a gene-blinded manner. Participant race and ethnicity were self-reported using fixed categories from the US Office of Management and Budget. Biological sex, race, and ethnicity were subsequently verified via genomic data. Methods of data collection included self-reported health and family histories, physical and neurological exams, standardized objective measurements for neuropsychology, cognitive behavioral screening, balance, bradykinesia, and companion/informant-, self-, and clinician-reported current behavioral, physical, and functional capacity assessment. Enrollment sites conduct the CADASIL standardized protocol at baseline and 18 and 36 months thereafter (**Table 1**).

**Table 1.**
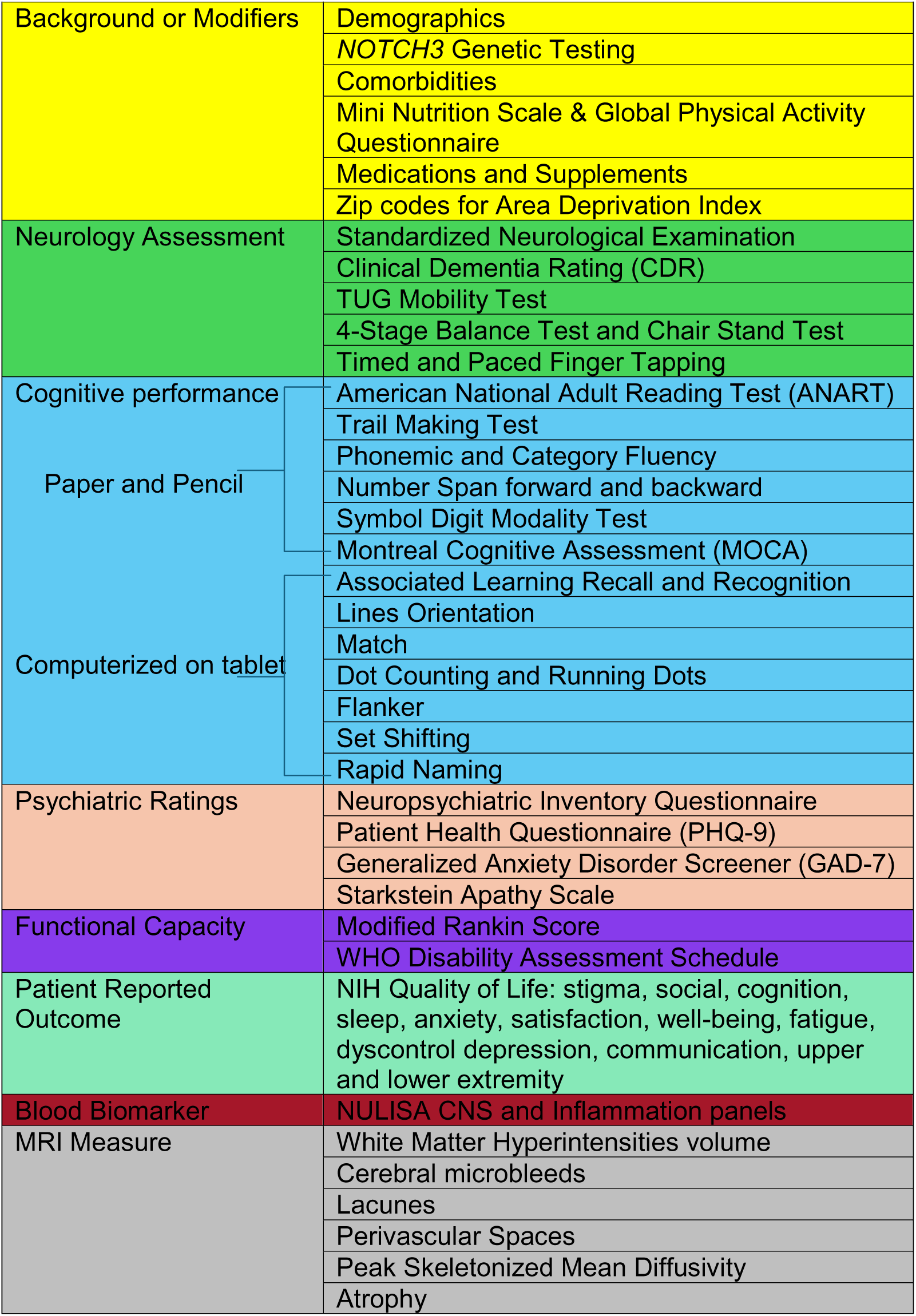
Standardized Protocol for the United States CADASIL Consortium.

### Participant Selection and Cohort Definitions

Eligible participants included individuals aged 18 years or older from families with a known or suspected pathogenic *NOTCH3* variant. Exclusion criteria were additional neurological diagnoses causing clinical findings to be uninterpretable, active uncontrolled systemic diseases, or moderate disability as defined by a modified Rankin Scale (mRS) score >3 (i.e., loss of unassisted walking). Companions/informants were enrolled, when possible, to provide external documentation of participants’ phenotypic changes.

Recruitment strategies included presentations at professional and community meetings, Webinars, collaborations with lay organizations (i.e., cureCADASIL, Youngtimers, Sisters’ Hope, United Leukodystrophy Foundation, The CADASIL Foundation), consultations with investigators from other studies of genetic neurodegenerative disease, and electronic medical record reviews.

### Multimodal Biomarker Assessments and Genetic Analyses

To minimize heterogeneity and align outcomes with Alzheimer’s Disease Research Center initiatives, MRI acquisition protocols were based on MarkVCID and ADNI3 frameworks, incorporating high-resolution 3D T1-weighted, 3D T2-weighted fluid-attenuated inversion recovery (FLAIR) diffusion, diffusion-weighted imaging (DWI), susceptibility-weighted quantitative susceptibility mapping (QSM), arterial spin labeling (ASL) perfusion, and resting-state fMRI sequences.^16,17^ All participants completed scans at 3T on Siemens Prisma or GE Discovery MR750 systems using multichannel head coils and site-specific software versions. Centralized quality assurance was conducted by trained personnel and neuroradiologists. Quality assurance involved automated screening for implausible or extreme values and manual visual inspection of segmentation masks for all flagged cases and a random subset of non-flagged cases. USCC imaging outcomes include white matter hyperintensity (WMH), cerebral microbleeds (CMB), lacunar infarcts, enlarged perivascular spaces (ePVS), cortical thickness, diffusion signatures of microstructural damage, structural and functional connectivity, and cerebral blood flow, in accordance with STRIVE neuroimaging standards.^18^

The National Centralized Repository for Alzheimer’s Disease and Related Dementias (NCRAD) processed and biobanked biospecimens,^20^ with central genetic testing conducted at the University of Wisconsin–Madison Next Generation Sequencing Division. DNA concentration was verified using the Qubit® dsDNA HS Assay Kit (Life Technologies, Grand Island, NY). Samples were prepared with the NEBNext Enzymatic Methyl-Seq v2 Kit (New England Biolabs). Quality and quantity of the finished libraries were assessed using an Agilent Tapestation (Agilent, Santa Clara, CA) and Qubit® dsDNA HS Assay Kit, respectively. Paired end, 150 bp sequencing was performed using the Illumina NovaSeq X Plus (Illumina, San Diego, CA). Plasma, RNA, buffy coat, and serum were obtained. Plasma proteomic profiling was performed at the UW Alzheimer’s Disease Research Center Biomarker Laboratory using the Nucleic acid-linked immunosandwich assay (NULISAseq™) CNS Disease Panel 120. NEFL assays were performed on the Alamar ARGO HT System according to the manufacturer’s protocol by laboratory personnel blinded to participant characteristics and imaging results. Counts for each analyte were first normalized to the internal control within each sample well. The internal-control-normalized counts were then divided by the analyte-specific median of the interplate control replicates on the same plate. The resulting values were rescaled and log2 transformed to generate NULISA Protein Quantification (NPQ) values.^22^ NPQ represents relative protein abundance. Values can be compared across participants for the same analyte; NPQ values from different analytes are not directly comparable.

To account for molecular heterogeneity, *NOTCH3* pathogenic or likely pathogenic variants were stratified into CADASIL low-, medium-, and high-risk tiers utilizing epidermal growth factor-like repeat (EGFr) domain location as designated by Hack et al.^8^ These tiers are determined by the *NOTCH3*^cys^ variant frequency odds ratio (NVFOR), which represents the odds that a *NOTCH3*^cys^ variant is located in a specific EGFr domain in CADASIL cohorts relative to the odds in *NOTCH3*^cys^-positive individuals from population databases. Risk tiers are defined as high-risk (HR) EGFr (NVFOR >5), medium-risk (MR) EGFr (NVFOR 0.20–5), and low-risk (LR) EGFr (NVFOR <0.20).

### Statistical Analysis

Continuous variables were summarized as medians with interquartile ranges (IQR). Categorical variables were summarized as frequencies and percentages. Percentages were calculated using non-missing observations for each variable.

Baseline demographic and clinical characteristics were compared with controls and across CADASIL risk tiers using Fisher exact tests for categorical variables and Kruskal-Wallis tests for continuous/ordinal variables. When the omnibus test was significant, post hoc pairwise comparisons were performed using exact logistic regression for categorical variables and the Dwass-Steel-Critchlow-Fligner procedure for continuous/ordinal variables.^19–21^

The current Area Deprivation Index (ADI),^22–24^ Mini Nutrition Assessment (MNA) Scale, Global Physical Activity Questionnaire (GPAQ), Clinical Dementia Rating (CDR), Montreal Cognitive Assessment (MoCA), modified Rankin Scale (mRS), and the World Health Organization Disability Assessment Schedule (WHODAS) were scored according to published guidelines.^25–31^ Summary scores were calculated only for participants with sufficient non-missing item-level data according to published guidelines.

Associations between participant characteristics and clinical outcomes were evaluated using univariable and multivariable general linear models. Candidate explanatory variables were specified *a priori* and included sex, race, Hispanic ethnicity, education (years), age (years) and risk tier. CDR Global Score was analyzed using univariable and multivariable ordinal logistic regression with a cumulative logit link. The proportional odds assumption was evaluated using a published score test.^32^ All statistical tests were two-sided. Statistical significance was defined as p < .05. Analyses used SAS version 9.4 (Cary, NC).

## Results

### Cohort Demographics and Genetic Variation

A total of 560 participants completed baseline visits. Of these, 46 participants exhibited variants of uncertain significance (VUS; 8.2%); 55 participants (9.8%) had pending genetic analyses. Excluding these individuals yielded an analytic subset of 459 participants who are reported in this manuscript (**Figure 1**): 343 persons with a CADASIL-causing *NOTCH3* variant (74.7%); 116 non-carrier family controls (25.3%).

The analytic cohort included 92 unique *NOTCH3* variants. The most common variant by unique participant count was located within EGFr domain 3 (n = 72), followed by domains 4 (n = 47), 5 (n = 37), 2 (n = 36), and 21 (n = 20). Concurrently, EGFr domain 21 represented the site with the highest number of unique variants (n = 14), followed by domains 1 (n = 13), 2 (n = 11), and 4.

Cases were stratified into LR (24/343 [7.0%]), MR (63/343 [18.4%]), and HR (256/343 [74.6%]) tiers (**Table 2**). The cohort had a median age of 47.9 years (IQR, 38.7–59.1), median education of 16.0 years (IQR, 14.0–17.0), and median predicted Wechsler Adult Intelligence Scale–Fourth Edition (WAIS-IV) Full Scale IQ of 111.7 (IQR, 105.9–116.6).^33^ Most participants were female (62.5%), White (91.4%), and non-Hispanic/Latino (91.5%).

**Table 2.** Participant and clinical characteristics by CADASIL risk tier.

|  | **CONTROL**<br>**(N=116)** | **LOW**<br>**(N=24)** | **MEDIUM**<br>**(N=63)** | **HIGH**<br>**(N=256)** | **Total**<br>**(N=459)** | **p** |
| --- | --- | --- | --- | --- | --- | --- |
| **Age** |  |  |  |  |  | .030<sup>1</sup> |
| N (Missing) | 116 (0) | 24 (0) | 63 (0) | 256 (0) | 459 (0) |  |
| Median (IQR) | 46.1 (33.0,<br>57.8) | 46.0 (39.4,<br>54.7) | 55.4 (43.2,<br>65.2) | 48.8 (38.8,<br>58.9) | 47.9 (38.7,<br>59.1) |  |
| **Age at diagnosis** |  |  |  |  |  | .025<sup>1</sup> |
| N (Missing) |  | 3 (21) | 19 (44) | 86 (170) | 108 (351) |  |
| Median (IQR) |  | 40.0 (36.0,<br>48.0) | 52.0 (45.0,<br>56.0) | 44.0 (37.0,<br>50.0) | 45.0 (37.5,<br>51.0) |  |
| **Sex, n (%)** |  |  |  |  |  | .718<sup>2</sup> |
| Female | 76 (65.5%) | 16 (66.7%) | 41 (65.1%) | 154 (60.2%) | 287 (62.5%) |  |
| Male | 40 (34.5%) | 8 (33.3%) | 22 (34.9%) | 102 (39.8%) | 172 (37.5%) |  |
| **Years of Education** |  |  |  |  |  | .064<sup>1</sup> |
| N (Missing) | 116 (0) | 24 (0) | 63 (0) | 256 (0) | 459 (0) |  |
| Median (IQR) | 16.0 (15.0,<br>18.0) | 15.5 (14.0,<br>17.0) | 16.0 (13.0,<br>18.0) | 16.0 (14.0,<br>16.0) | 16.0 (14.0,<br>17.0) |  |
| **Estimated IQ (ANART)** |  |  |  |  |  | .049<sup>1</sup> |
| N (Missing) | 115 (1) | 22 (2) | 61 (2) | 244 (12) | 442 (17) |  |
| Median (IQR) | 112.7 (107.8,<br>117.6) | 110.3 (104.9,<br>115.7) | 113.7 (106.9,<br>118.6) | 111.7 (104.9,<br>115.7) | 111.7 (105.9,<br>116.6) |  |
| **Race, n (%)** |  |  |  |  |  | .018<sup>2</sup> |
| non-White | 7 (6.1%) | 6 (25.0%) | 8 (12.9%) | 18 (7.1%) | 39 (8.6%) |  |
| White | 108 (93.9%) | 18 (75.0%) | 54 (87.1%) | 234 (92.9%) | 414 (91.4%) |  |
| Missing | 1 | 0 | 1 | 4 | 6 |  |
| **Ethnicity, n (%)** |  |  |  |  |  | .002<sup>2</sup> |
| Not Hispanic or Latino | 113 (97.4%) | 21 (87.5%) | 51 (81.0%) | 235 (91.8%) | 420 (91.5%) |  |
| Hispanic or Latino | 3 (2.6%) | 3 (12.5%) | 12 (19.0%) | 21 (8.2%) | 39 (8.5%) |  |
| **Stroke, n (%)** |  |  |  |  |  | <.001<sup>2</sup> |
| No | 114 (98.3%) | 19 (79.2%) | 43 (69.4%) | 146 (57.9%) | 322 (70.9%) |  |
| Yes | 2 (1.7%) | 5 (20.8%) | 19 (30.6%) | 106 (42.1%) | 132 (29.1%) |  |
| Missing or unknown | 0 | 0 | 1 | 4 | 5 |  |
| **TIA or ministroke, n (%)** |  |  |  |  |  | <.001<sup>2</sup> |
| No | 111 (95.7%) | 17 (70.8%) | 42 (66.7%) | 156 (62.2%) | 326 (71.8%) |  |
| Yes | 5 (4.3%) | 7 (29.2%) | 21 (33.3%) | 95 (37.8%) | 128 (28.2%) |  |
| Missing or unknown | 0 | 0 | 0 | 5 | 5 |  |
| **Age at first stroke or TIA** |  |  |  |  |  | .527<sup>1</sup> |
| N (Missing or not applicable) | 6 (110) | 9 (15) | 25 (38) | 149 (107) | 189 (270) |  |
| Median (IQR) | 48.0 (41.0, 53.0) | 43.0 (36.0, 45.0) | 48.0 (35.0, 53.0) | 43.0 (35.0, 50.0) | 43.0 (35.0, 51.0) |  |
| **Migraine, n (%)** |  |  |  |  |  | .002<sup>2</sup> |
| No | 47 (40.5%) | 12 (50.0%) | 21 (33.3%) | 62 (24.3%) | 142 (31.0%) |  |
| Yes | 69 (59.5%) | 12 (50.0%) | 42 (66.7%) | 193 (75.7%) | 316 (69.0%) |  |
| Missing | 0 | 0 | 0 | 1 | 1 |  |
| **Migraine with aura, n (%)** |  |  |  |  |  | .001<sup>2</sup> |
| No | 34 (49.3%) | 5 (41.7%) | 15 (35.7%) | 46 (24.0%) | 100 (31.7%) |  |
| Yes | 35 (50.7%) | 7 (58.3%) | 27 (64.3%) | 146 (76.0%) | 215 (68.3%) |  |
| Missing or not applicable | 47 | 12 | 21 | 64 | 144 |  |

|  | **CONTROL**<br>**(N=116)** | **LOW**<br>**(N=24)** | **MEDIUM**<br>**(N=63)** | **HIGH**<br>**(N=256)** | **Total**<br>**(N=459)** | ***p*** |
| --- | --- | --- | --- | --- | --- | --- |
| **Mild Cognitive Impairment, n (%)** |  |  |  |  |  | **<.001<sup>2</sup>** |
| No | 116 (100.0%) | 23 (95.8%) | 55 (87.3%) | 230 (90.2%) | 424 (92.6%) |  |
| Yes | 0 (0.0%) | 1 (4.2%) | 8 (12.7%) | 25 (9.8%) | 34 (7.4%) |  |
| Missing | 0 | 0 | 0 | 1 | 1 |  |
| **Dementia, n (%)** |  |  |  |  |  | **.202<sup>2</sup>** |
| No | 116 (100.0%) | 24 (100.0%) | 61 (96.8%) | 247 (96.9%) | 448 (97.8%) |  |
| Yes | 0 (0.0%) | 0 (0.0%) | 2 (3.2%) | 8 (3.1%) | 10 (2.2%) |  |
| Missing | 0 | 0 | 0 | 1 | 1 |  |
| **Count of neurological abnormalities** |  |  |  |  |  | **<.001<sup>1</sup>** |
| N (Missing) | 116 (0) | 23 (1) | 60 (3) | 246 (10) | 445 (14) |  |
| Median (IQR) | 1.0 (0.0, 2.0) | 0.0 (0.0, 2.0) | 1.5 (1.0, 3.0) | 1.0 (0.0, 3.0) | 1.0 (0.0, 2.0) |  |
| **Count of neurological abnormalities (recoded), n (%)** |  |  |  |  |  | **<.001<sup>1</sup>** |
| None | 53 (45.7%) | 13 (56.5%) | 14 (23.3%) | 72 (29.3%) | 152 (34.2%) |  |
| One | 31 (26.7%) | 2 (8.7%) | 16 (26.7%) | 52 (21.1%) | 101 (22.7%) |  |
| Two | 24 (20.7%) | 3 (13.0%) | 12 (20.0%) | 43 (17.5%) | 82 (18.4%) |  |
| Three or more | 8 (6.9%) | 5 (21.7%) | 18 (30.0%) | 79 (32.1%) | 110 (24.7%) |  |
| Missing | 0 | 1 | 3 | 10 | 14 |  |
| **Patient Health Questionnaire-9 (estimated)** |  |  |  |  |  | **.012<sup>1</sup>** |
| N (Missing) | 113 (3) | 24 (0) | 62 (1) | 251 (5) | 450 (9) |  |
| Median (IQR) | 2.0 (0.0, 5.0) | 3.5 (0.0, 6.5) | 5.0 (0.0, 7.0) | 3.0 (0.0, 8.0) | 3.0 (0.0, 7.0) |  |
| **Generalized Anxiety Disorder-7 (estimated)** |  |  |  |  |  | .009<sup>1</sup> |
| N (Missing) | 113 (3) | 24 (0) | 61 (2) | 251 (5) | 449 (10) |  |
| Median (IQR) | 4.0 (2.0, 7.0) | 7.0 (3.5, 10.0) | 6.0 (2.0, 10.0) | 5.0 (2.0, 9.0) | 5.0 (2.0, 9.0) |  |
| **Charlson Comorbidity Index (estimated)** |  |  |  |  |  | .479<sup>1</sup> |
| N (Missing) | 116 (0) | 24 (0) | 63 (0) | 256 (0) | 459 (0) |  |
| Median (IQR) | 2.0 (0.5, 2.0) | 1.0 (0.0, 2.0) | 2.0 (1.0, 3.0) | 2.0 (1.0, 3.0) | 2.0 (1.0, 3.0) |  |
| **Hypertension, n (%)** |  |  |  |  |  | .610<sup>2</sup> |
| No | 86 (74.1%) | 19 (79.2%) | 48 (76.2%) | 204 (80.0%) | 357 (77.9%) |  |
| Yes | 30 (25.9%) | 5 (20.8%) | 15 (23.8%) | 51 (20.0%) | 101 (22.1%) |  |
| Missing | 0 | 0 | 0 | 1 | 1 |  |
| **Diabetes, n (%)** |  |  |  |  |  | .047<sup>2</sup> |
| No | 109 (94.0%) | 21 (87.5%) | 54 (85.7%) | 242 (94.9%) | 426 (93.0%) |  |
| Yes | 7 (6.0%) | 3 (12.5%) | 9 (14.3%) | 13 (5.1%) | 32 (7.0%) |  |
| Missing | 0 | 0 | 0 | 1 | 1 |  |
| **Dyslipidemia, n (%)** |  |  |  |  |  | .011<sup>2</sup> |
| No | 82 (70.7%) | 13 (54.2%) | 29 (46.0%) | 154 (60.4%) | 278 (60.7%) |  |
| Yes | 34 (29.3%) | 11 (45.8%) | 34 (54.0%) | 101 (39.6%) | 180 (39.3%) |  |
| Missing | 0 | 0 | 0 | 1 | 1 |  |
| **Body Mass Index (kg/m)** |  |  |  |  |  | .426<sup>1</sup> |
| N (Missing) | 116 (0) | 23 (1) | 63 (0) | 255 (1) | 457 (2) |  |
| Median (IQR) | 27.8 (24.6, 32.1) | 30.5 (25.3, 35.0) | 29.2 (24.9, 36.2) | 28.7 (24.7, 33.0) | 28.4 (24.8, 33.0) |  |
| **Tobacco, n (%)** |  |  |  |  |  | .238<sup>2</sup> |
| Never | 63 (54.3%) | 15 (62.5%) | 35 (55.6%) | 138 (54.1%) | 251 (54.8%) |  |
| Former | 37 (31.9%) | 7 (29.2%) | 23 (36.5%) | 102 (40.0%) | 169 (36.9%) |  |
| Current | 16 (13.8%) | 2 (8.3%) | 5 (7.9%) | 15 (5.9%) | 38 (8.3%) |  |
| Missing | 0 | 0 | 0 | 1 | 1 |  |
| **Marijuana, n (%)** |  |  |  |  |  | <.001<sup>2</sup> |
| No | 34 (29.6%) | 15 (62.5%) | 29 (46.8%) | 132 (51.8%) | 210 (46.1%) |  |
| Yes | 81 (70.4%) | 9 (37.5%) | 33 (53.2%) | 123 (48.2%) | 246 (53.9%) |  |
| Missing | 1 | 0 | 1 | 1 | 3 |  |
| **MNA Total Score** |  |  |  |  |  | <.001<sup>1</sup> |
| N (Missing) | 114 (2) | 24 (0) | 61 (2) | 250 (6) | 449 (10) |  |
| Median (IQR) | 27.0 (25.5,<br>28.5) | 26.8 (25.5,<br>27.8) | 26.0 (23.0,<br>27.0) | 26.0 (24.0,<br>27.5) | 26.5 (24.0,<br>28.0) |  |
| **MNA Nutritional Status, n (%)** |  |  |  |  |  | .001<sup>1</sup> |
| Normal nutritional status | 100 (87.7%) | 21 (87.5%) | 39 (63.9%) | 188 (75.2%) | 348 (77.5%) |  |
| At risk of malnutrition | 13 (11.4%) | 3 (12.5%) | 19 (31.1%) | 56 (22.4%) | 91 (20.3%) |  |
| Malnourished | 1 (0.9%) | 0 (0.0%) | 3 (4.9%) | 6 (2.4%) | 10 (2.2%) |  |
| Missing | 2 | 0 | 2 | 6 | 10 |  |
| **Total physical activity**<br>**(min/day)** |  |  |  |  |  | .003<sup>1</sup> |
| N (Missing) | 114 (2) | 24 (0) | 62 (1) | 251 (5) | 451 (8) |  |
| Median (IQR) | 73.9 (31.4,<br>180.0) | 34.3 (10.7,<br>76.1) | 47.9 (11.4,<br>107.1) | 42.9 (17.1,<br>111.4) | 51.4 (17.1,<br>120.0) |  |
| **Meets WHO physical activity recommendation, n (%)** |  |  |  |  |  | **.003<sup>2</sup>** |
| No | 15 (13.2%) | 9 (37.5%) | 20 (32.3%) | 69 (27.5%) | 113 (25.1%) |  |
| Yes | 99 (86.8%) | 15 (62.5%) | 42 (67.7%) | 182 (72.5%) | 338 (74.9%) |  |
| Missing | 2 | 0 | 1 | 5 | 8 |  |
| **WHODAS Total Score** |  |  |  |  |  | **<.001<sup>1</sup>** |
| N (Missing) | 113 (3) | 24 (0) | 62 (1) | 249 (7) | 448 (11) |  |
| Median (IQR) | 1.0 (0.0, 4.0) | 3.5 (0.0, 10.5) | 6.0 (1.0, 13.0) | 4.0 (1.0, 11.0) | 3.0 (1.0, 10.0) |  |
| **CDR Box Sum** |  |  |  |  |  | **<.001<sup>1</sup>** |
| N (Missing) | 108 (8) | 24 (0) | 60 (3) | 239 (17) | 431 (28) |  |
| Median (IQR) | 0.0 (0.0, 0.0) | 0.5 (0.0, 1.3) | 0.5 (0.0, 1.5) | 0.5 (0.0, 2.5) | 0.0 (0.0, 1.5) |  |
| **CDR Global Score, n (%)** |  |  |  |  |  | **<.001<sup>1</sup>** |
| Normal | 87 (80.6%) | 11 (45.8%) | 31 (51.7%) | 107 (44.8%) | 236 (54.8%) |  |
| Very mild dementia | 21 (19.4%) | 13 (54.2%) | 29 (48.3%) | 110 (46.0%) | 173 (40.1%) |  |
| Mild dementia | 0 (0.0%) | 0 (0.0%) | 0 (0.0%) | 18 (7.5%) | 18 (4.2%) |  |
| Moderate dementia | 0 (0.0%) | 0 (0.0%) | 0 (0.0%) | 3 (1.3%) | 3 (0.7%) |  |
| Severe dementia | 0 (0.0%) | 0 (0.0%) | 0 (0.0%) | 1 (0.4%) | 1 (0.2%) |  |
| Missing | 8 | 0 | 3 | 17 | 28 |  |
| **MoCA Total Score** |  |  |  |  |  | **.012<sup>1</sup>** |
| N (Missing) | 116 (0) | 24 (0) | 62 (1) | 253 (3) | 455 (4) |  |
| Median (IQR) | 27.0 (25.0, 29.0) | 27.0 (25.5, 28.0) | 27.0 (24.0, 28.0) | 26.0 (24.0, 28.0) | 27.0 (25.0, 28.0) |  |
| **MoCA Total Score (recoded), n (%)** |  |  |  |  |  | **.035<sup>2</sup>** |
| < 26 Points | 31 (26.7%) | 6 (25.0%) | 26 (41.9%) | 102 (40.3%) | 165 (36.3%) |  |
| >= 26 Points | 85 (73.3%) | 18 (75.0%) | 36 (58.1%) | 151 (59.7%) | 290 (63.7%) |  |
| Missing | 0 | 0 | 1 | 3 | 4 |  |
| **Modified Rankin Scale, n (%)** |  |  |  |  |  | **<.001<sup>1</sup>** |
| No symptoms | 86 (74.1%) | 12 (50.0%) | 23 (36.5%) | 82 (32.0%) | 203 (44.2%) |  |
| No significant disability | 25 (21.6%) | 7 (29.2%) | 19 (30.2%) | 97 (37.9%) | 148 (32.2%) |  |
| Slight disability | 2 (1.7%) | 3 (12.5%) | 16 (25.4%) | 40 (15.6%) | 61 (13.3%) |  |
| Moderate disability | 0 (0.0%) | 1 (4.2%) | 3 (4.8%) | 21 (8.2%) | 25 (5.4%) |  |
| mRS < 4; exact score unavailable<sup>a</sup> | 3 (2.6%) | 1 (4.2%) | 2 (3.2%) | 16 (6.3%) | 22 (4.8%) |  |
| **White Matter Hyperintensity (WMH) burden, n (%)** |  |  |  |  |  | **<.001<sup>1</sup>** |
| Zero | 0 (0.0%) | 0 (0.0%) | 1 (1.9%) | 0 (0.0%) | 1 (0.2%) |  |
| Mild | 101 (96.2%) | 14 (77.8%) | 23 (42.6%) | 61 (26.1%) | 199 (48.4%) |  |
| Moderate | 3 (2.9%) | 3 (16.7%) | 12 (22.2%) | 89 (38.0%) | 107 (26.0%) |  |
| Severe | 1 (1.0%) | 1 (5.6%) | 18 (33.3%) | 84 (35.9%) | 104 (25.3%) |  |
| Missing | 11 | 6 | 9 | 22 | 48 |  |
| **Lacune burden, n (%)** |  |  |  |  |  | **<.001<sup>1</sup>** |
| Zero | 17 (16.3%) | 4 (23.5%) | 4 (7.5%) | 13 (5.6%) | 38 (9.4%) |  |
| Single | 31 (29.8%) | 3 (17.6%) | 8 (15.1%) | 25 (10.8%) | 67 (16.5%) |  |
| Few | 45 (43.3%) | 9 (52.9%) | 26 (49.1%) | 85 (36.8%) | 165 (40.7%) |  |
| Multiple | 11 (10.6%) | 1 (5.9%) | 15 (28.3%) | 108 (46.8%) | 135 (33.3%) |  |
| Missing | 12 | 7 | 10 | 25 | 54 |  |
| **Perivascular-space (PVS) burden, n (%)** |  |  |  |  |  | **<.001<sup>1</sup>** |
| Mild | 73 (69.5%) | 11 (61.1%) | 13 (24.5%) | 75 (32.1%) | 172 (42.0%) |  |
| Moderate | 20 (19.0%) | 4 (22.2%) | 18 (34.0%) | 80 (34.2%) | 122 (29.8%) |  |
| Severe | 12 (11.4%) | 3 (16.7%) | 22 (41.5%) | 79 (33.8%) | 116 (28.3%) |  |
| Missing | 11 | 6 | 10 | 22 | 49 |  |
| **Cerebral Microbleed (CMB)**<br>**burden, n (%)** |  |  |  |  |  | **<.001<sup>1</sup>** |
| Zero | 72 (73.5%) | 11 (78.6%) | 29 (56.9%) | 89 (40.8%) | 201 (52.8%) |  |
| Single | 17 (17.3%) | 2 (14.3%) | 6 (11.8%) | 39 (17.9%) | 64 (16.8%) |  |
| Few | 9 (9.2%) | 1 (7.1%) | 13 (25.5%) | 47 (21.6%) | 70 (18.4%) |  |
| Multiple | 0 (0.0%) | 0 (0.0%) | 3 (5.9%) | 43 (19.7%) | 46 (12.1%) |  |
| Missing | 18 | 10 | 12 | 38 | 78 |  |
| **NfL (pg/mL)** |  |  |  |  |  | **<.001<sup>1</sup>** |
| N (Missing) | 116 (0) | 24 (0) | 63 (0) | 255 (1) | 458 (1) |  |
| Median (IQR) | 10.5 (10.0,<br>11.1) | 10.4 (10.1,<br>10.9) | 10.9 (10.3,<br>11.5) | 10.8 (10.2,<br>11.8) | 10.7 (10.2,<br>11.5) |  |
| <sup>1</sup>Kruskal-Wallis p-value; <sup>2</sup>Fisher Exact p-value; ANART = American National Adult Reading Test; TIA = Transient Ischemic Attack; MNA = Mini Nutritional Assessment; WHO = World Health Organization; CDR = Clinical Dementia Rating; MoCA = Montreal Cognitive Assessment; WHODAS = World Health Organization Disability Assessment Schedule; NfL = Neurofilament light. <sup>a</sup>For the modified Rankin Scale (mRS), exact scores were unavailable for 22 participants; these participants were known only to have an mRS < 4 and are shown descriptively but were excluded from the hypothesis test for mRS. |  |  |  |  |  |  |

Demographic characteristics differed across CADASIL risk tiers. Post hoc testing showed LR cases were more frequently non-White than controls (*p* = .02) or HR cases (*p* = .02). MR cases were older than controls (*p* = .03) and more likely to identify as Hispanic/Latino than controls (*p* = .001) or HR cases (*p* = .03). Sex (*p* = .72) and educational attainment (*p* = .06) did not differ significantly across case groups and controls.

### Clinical Manifestations and Risk Factors

Self-reported stroke and TIA were significantly more prevalent across all CADASIL risk tiers than controls (all *p* < .05). Ischemic stroke prevalence was highest in HR cases (42.1%). Among individuals experiencing ischemic events, age at first stroke or TIA did not differ by risk tier (*p* =.53).

Migraine was more prevalent in the HR tier than in both controls (75.7% vs 59.5%; *p* = .003) and the LR tier (75.7% vs 50.0%; *p* = .02). Among those with migraines, HR cases had significantly higher rates of migraine with aura than controls (76.0% vs 50.7%; *p* < .001). Relative to controls, mild cognitive impairment was also more common in the MR (12.7% vs 0.0%; *p* < .001) and HR (9.8% vs 0.0%; *p* < .001) tiers. Dementia diagnosis did not differ among groups (*p* = .20). However, the count of neurological abnormalities was significantly higher in the medium-(*Mdn* = 1.5, IQR: 1.0 – 3.0; *p* = .001) and HR (*Mdn* = 1.0, IQR: 0.0 – 3.0; *p* < .001) tiers relative to controls (*Mdn* = 1.0, IQR: 0.0 – 2.0). Patient Health Questionnaire (PHQ9) showed that mood was worse in MR and HR groups than in controls or LR tiers. Anxiety assessed with the Generalized Anxiety Disorder-7-item (GAD-7) screening scale was higher in the MR group.

Charleson Comordibity Indices (CCI) did not differ at baseline between case groups and controls.^34^ The baseline prevalence of hypertension was similar across risk tiers (*p* = .61). Diabetes prevalence was, however, significantly higher in the MR tier than in the HR tier (14.3% vs 5.1%; *p* = .03). Dyslipidemia was significantly more common in the MR tier than controls (54.0% vs 29.3%; *p* = .002). Neither use of tobacco (*p* = .24) nor body mass index (*p* = .43) were different among groups. Marijuana use was greater in controls (70.4%) than the LR (37.5%; *p* = .005), MR (53.2%; *p* = .04), and HR (48.2%; *p* < .001) tiers.

MNA total scores differed significantly across groups (*p* < .001). Compared to controls, scores were significantly lower in the HR (*Mdn* = 26.0 vs. 27.0; *p* < .001) and MR (*Mdn* = 26.0 vs. 27.0; *p* = .004) tiers. MNA nutritional status also differed significantly across groups (*p* = .001), with poorer nutritional status in the HR (*z* = 2.7; *p* = .03) and MR (*z* = 3.7; *p* = .001) tiers compared with controls. The proportion of participants at risk of malnutrition was 22.4% in the HR and 31.1% in the MR tiers, compared with 11.4% among controls.

Total physical activity, as measured by the GPAQ, also differed significantly across groups (*p* = .003). Participants’ total physical activity was significantly lower in the HR (*Mdn* = 42.9 vs. 73.9 min/day; *p* = .01) and LR (*Mdn* = 34.3 vs. 73.9 min/day; *p* = .02) tiers compared with controls. In fact, the proportion of participants meeting WHO physical activity recommendations was also lower in the HR (72.5% vs. 86.8%; *p* = .003), MR (67.7% vs. 86.8%; *p* = .005), and LR (62.5% vs. 86.8%; *p* = .02) tiers compared with controls. The WHO Disability Assessment Schedule total score was worse in HR and HR tiers (all p<.001) compared with controls.

### Clinical Dementia Rating, MOCA and Functional Outcomes

The CDR Box Sum was significantly higher in the HR (*Mdn* = 0.5 vs. 0.0; *p* < .001), MR (*Mdn* = 0.5 vs. 0.0; *p* < .001), and LR (*Mdn* = 0.5 vs. 0.0; *p* = .003) tiers compared with controls. On CDR, HR cases showed significant impairment relative to controls across 8 of the 9 domains (all *p* < .05). MR cases displayed widespread deficits whereas LR cases showed more restricted domain-specific impairments in memory, judgment and problem solving, community affairs, and home and hobbies (all *p* < .05; **Supplemental Table S1**).

On multivariable analysis adjusting for age, sex, race, and educational attainment, CADASIL risk tier remained associated with worse CDR Global Scores (*p* < .001; **Table 3**). Compared with controls, the odds of classification into a more impaired CDR category were higher for the LR (*OR =* 4.31, 95% CI: 1.64 – 11.34; *p* = .003), MR (*OR* = 2.55, 95% CI: 1.22 – 5.33; *p* = .01), and HR (*OR* = 5.08, 95% CI: 2.87 – 8.99; *p* < .001) tiers.

**Table 3.** Odds of Worse Clinical Dementia Rating Global Scores by CADASIL Risk Tier and Participant Characteristics.

|  | **Valid N** | **Unadjusted OR**<br>**(95% CI)** | ***p*** | **Adjusted OR**<br>**(95% CI)** | ***p*** |
| --- | --- | --- | --- | --- | --- |
| **Group (omnibus)** | 431 |  | <.001<sup>a</sup> |  | <.001<sup>a</sup> |
| High vs Control |  | 5.50 (3.20–9.44) | <.001 | 5.08 (2.87–8.99) | <.001 |
| Medium vs Control |  | 3.60 (1.80–7.19) | <.001 | 2.55 (1.22–5.33) | .01 |
| Low vs Control |  | 4.41 (1.76–11.01) | .002 | 4.31 (1.64–11.34) | .003 |
| **Male vs Female** | 431 | 1.37 (0.93–2.01) | .11 | 1.52 (0.99–2.31) | .054 |
| **Non-White vs White** | 425 | 1.19 (0.62–2.28) | .61 | 1.70 (0.82–3.49) | .15 |
| **Hispanic vs Non-Hispanic** | 431 | 1.42 (0.74–2.73) | .30 |  |  |
| **Education (per year)** | 431 | 0.84 (0.77–0.91) | <.001 | 0.83 (0.76–0.91) | <.001 |
| **Age (per year)** | 431 | 1.04 (1.03–1.06) | <.001 | 1.05 (1.03–1.07) | <.001 |
| Valid N = The number of observations used to compute the unadjusted estimate. The number of observations used to compute the adjusted estimates = 425. Abbreviations: CDR = Clinical Dementia Rating; OR = odds ratio; CI = confidence interval. <sup>a</sup>Omnibus *p*-value. |  |  |  |  |  |

On univariable analysis, participants in the HR tier had a lower MoCA visuospatial/executive sub-score (*p* < .001) and lower total scores (*p* = .01) than controls. Although the omnibus test for MoCA delayed recall was significant (*p* = .04), no post hoc pairwise comparisons were significant. After adjusting for age, sex, race, and years of education, there were no significant differences in MoCA total scores among the groups (*p* = .07; **Table 4**).

**Table 4.** MOCA total score as a function of cohort status and participant characteristics.

|  | **Valid**<br>**N** | **Unadjusted**<br>**$\hat{\beta}$ (95% CI)** |  | **Adjusted**<br>**$\hat{\beta}$ (95% CI)** |  |
| --- | --- | --- | --- | --- | --- |
| **Group** | 455 |  | .01<sup>a</sup> |  | .07<sup>a</sup> |
| High vs Control |  | -1.23 (-1.96 to -0.51) | .001 | -0.65 (-1.30 to -0.01) | .048 |
| Medium vs Control |  | -1.00 (-2.02 to 0.02) | .055 | 0.01 (-0.91 to 0.93) | .98 |
| Low vs Control |  | -0.03 (-1.48 to 1.43) | .97 | 0.46 (-0.83 to 1.75) | .48 |
| **Sex:** Male vs Female | 455 | -0.56 (-1.20 to 0.07) | .08 | -0.75 (-1.32 to -0.20) | .01 |
| **Race:** non-White vs White | 449 | -0.65 (-1.76 to 0.45) | .25 | -1.29 (-2.27 to -0.31) | .01 |
| **Hispanic:** Yes vs No | 455 | -1.83 (-2.92 to -0.74) | .001 |  |  |
| **Education** (per year) | 455 | 0.43 (0.30 to 0.55) | <.001 | 0.45 (0.33 to 0.56) | <.001 |
| **Age** (per year) | 455 | -0.08 (-0.11 to -0.06) | <.001 | -0.09 (-0.11 to -0.07) | <.001 |
| **Note:** Valid N = The number of observations used for the unadjusted estimates. The number of observations used for the adjusted estimates = 449. <sup>a</sup>Omnibus *p*-value. |  |  |  |  |  |

Functional impairment differed across risk tiers. Compared with controls, all cases demonstrated significantly worse mRS scores (all *p* < .05). Additionally, compared to controls, disability burden and functional dependence were greater in the MR and HR tiers, which had higher WHODAS-12 scores (both *p* < .001).

### Biospecimen and Neuroimaging

Plasma was available for 99.6% of the cohort, followed by RNA (99.2%), buffy coat (98.8%), and serum (97.8%). NULISA-measured neurofilament light chain (NfL) differed across risk tiers (*p* < .001), with higher levels in the MR (*z* = 2.86, *p* = .02) and HR (*z* = 3.96, *p* < .001) tiers compared with controls; the LR tier did not differ from controls (*z* = −0.03, *p* = .99; **Table 2**).

Completed anatomical MRI (T1w/T2w/FLAIR) was captured in 89.1% of participants, DWI in 82.6%, QSM in 87.5%, ASL in 86.5%, and resting-state fMRI in 82.4%. WMH, lacune, ePVS), and CMB burden each differed significantly across risk tiers (all *p* < .05). Compared with controls, the HR tier had greater WMH (*z* = 11.27, *p* < .001), lacune (*z* = 7.16, *p* < .001), ePVS (*z* = 6.31, *p* < .001), and CMB (*z* = 6.15, *p* < .001) burden. The MR tier also had greater WMH (*z* = 7.23, *p* < .001), lacune (*z* = 3.35, *p* = .005), ePVS (*z* = 5.56, *p* < .001), and CMB (*z* = 2.60, *p* = .045) burden when compared with controls. In contrast, the LR tier differed from controls only in WMH burden (*z* = 2.91, *p* = .02), with no differences in lacune (*z* = −0.15, *p* = .99), ePVS (*z* = 0.74, *p* = .88), or CMB (*z* = −0.41, *p* = .98) burden.

Among risk tiers, HR participants had greater WMH (*z* = 4.17, *p* < .001), lacune (*z* = 3.63, *p* = .002), and CMB (*z* = 2.89, *p* = .02) burden than those in the LR tier while PVS burden was greater in the MR than LR tiers (*z* = 2.69, *p* = .04). No imaging burden measures differed significantly between the MR and HR tiers (all *p* > .05).

## Discussion

The USCC establishes a national, multisite platform for investigating the clinical heterogeneity of *NOTCH3*-associated SVD. This baseline analysis included 343 participants with pathogenic or likely *NOTCH3* variants and 116 non-carriers recruited from affected families. Cases demonstrated greater cognitive and functional burden than controls across risk tiers, but the pattern of differences varied by outcome. The HR group had the highest observed prevalence of ischemic stroke and migraine while the MR group had a greater burden of specific risk factors. Even participants in the LR tier showed differences from controls in everyday cognitive functioning and disability. These findings document clinically relevant variation within a cohort selected to include individuals without advanced disability and support the need to examine disease expression across multiple domains.

The USCC builds on earlier US studies of CADASIL severity and individual clinical manifestations. Anisetti and colleagues developed a clinical grading system from a multicenter sample, organizing participants according to manifestations ranging from migraine to ischemic events, cognitive impairment, and dependence on walking assistance.^35^ While smaller investigations examined relationships among migraine, circulating biomarkers, and MRI findings, Pan and colleagues used electronic health records to characterize cerebrovascular disease burden.^36,37^ The USCC extends this foundation through prospective assessments at 12 enrollment sites under a common protocol. Centralized genetic characterization and biospecimen collection accompany standardized MRI acquisition, detailed cognitive testing, and measures of daily functioning. Inclusion of non-carrier family controls provides an internal comparison group that shares aspects of familial background and experience with CADASIL.

European cohorts provide an important context for interpreting the genetic composition and clinical findings of the USCC. The United Kingdom national cohort included 485 genetically confirmed participants, with more than 80% carrying variants in EGFr domains 1–6. Proximal variants were associated with earlier stroke onset after adjustment for cardiovascular risk factors.^38^ Dutch and European investigations also established associations between variant position and disease severity, forming the basis for subsequent refinement of genetic risk classifications.^8,9^ The USCC applies the three-tier classification to a US cohort with broad representation of *NOTCH3* variants. Because this classification incorporates risk differences beyond the earlier EGFr 1–6 versus 7–34 division, its categories should be distinguished from the proximal–distal groupings used in many previous studies.

The baseline findings did not show a uniformly ordered clinical gradient across risk tiers. Although HR participants had substantial neurological and functional burden, the MR group had higher frequencies of diabetes, dyslipidemia, and psychiatric symptoms in selected comparisons. All three case groups had greater adjusted odds of classification into a more impaired CDR category than controls. These case–control associations do not establish that the tiers differ significantly from one another, and the small LR group limits precision. Similarly, the absence of a difference in age at first ischemic event among participants who had experienced an event does not establish equivalent stroke risk across tiers. That comparison excludes participants who remained event-free. Longitudinal follow-up and time-to-event analyses will be needed to evaluate age-dependent risk and the prognostic value of the classification.

Regional differences in variant distribution also influence comparisons with East Asian cohorts. In Taiwan, p.R544C accounts for a large majority of CADASIL pedigrees in some published series. Korean multicenter studies have reported a prominence of exon 11 variants (corresponding to the EGFr 13/14 interdomain boundary) and a more even distribution across proximal and distal EGFr domains than European referral cohorts.^39,40^ Pooled analyses have found more frequent reports of ischemic events and cognitive impairment in Asian samples, with migraine reported more frequently in predominantly European samples.^41^ Migraine prevalence in the USCC HR group, 75.7%, was similar to the 75.3% reported in a symptomatic UK referral cohort.^42^ Interpretation requires attention to diagnostic definitions, sex distribution, participant age, and recruitment methods.

Ascertainment is central to understanding these international differences. Stroke-service recruitment preferentially identifies participants who have already experienced a vascular event. Recruitment through affected families and community organizations can reach individuals earlier in their clinical course. The USCC also excluded participants with mRS scores >3, restricting representation of advanced disability or disease. Consequently, the reported lower frequency of severe manifestations cannot establish a milder US phenotype. Geographic comparisons will require harmonized definitions and adjustment for clinical stage and variant composition. The predominantly White composition of the present cohort further limits conclusions about ancestry-related differences within the US.

The distinction between genetic status and symptomatic disease is important throughout this work. A pathogenic or likely pathogenic *NOTCH3* variant establishes a molecular diagnosis, but it does not by itself specify whether an individual currently has attributable symptoms or functional impairment. MRI abnormalities can precede clinical manifestations. Genotype-first studies in UK Biobank and Geisinger have identified carriers with a broad range of expression, including individuals with limited clinical disease into later adulthood.^12,43^ These observations support separate characterization of molecular status, imaging burden, and clinical expression. The USCC permits these dimensions to be examined together although the proportion meeting a defined presymptomatic state will require explicit classification.

The cognitive findings illustrate the value of examining performance alongside everyday functioning. CADASIL risk tier remained associated with CDR Global Score after demographic adjustment whereas the adjusted association with total MoCA score did not reach statistical significance. While the CDR incorporates information about cognitive changes affecting daily activities, the MoCA provides a brief assessment of performance. Differences in these results may arise from the constructs assessed, score distributions, or the cohort’s high educational attainment. They do not establish that one instrument is more sensitive than the other. Detailed neuropsychological assessment and repeated measurement will help determine which outcomes detect meaningful change. Greater disability on the mRS and, in the MR and HR tiers, the WHODAS also demonstrates that functional burden is present among participants who remain able to walk independently.

The inclusion of nutrition and physical activity expands the characterization of CADASIL beyond manifestations traditionally reported in clinical registries. MR and HR participants had poorer nutritional status than controls, and participants in each risk tier were less likely to meet physical activity recommendations. These associations identify questions for longitudinal investigation. Reduced activity or nutritional risk could contribute to poorer outcomes, but emerging neurological symptoms and disability could also affect these behaviors. Baseline data cannot determine the direction of these relationships. Repeated assessments will allow evaluation of whether these measures predict subsequent change after accounting for initial disease burden.

A genetically defined cohort offers a useful approach to understanding mechanisms relevant to VCID. In CADASIL, investigators can begin with an established cause of vascular disease and examine why its expression varies among individuals. Hypertension, other genetic influences, and age-related pathology remain important. Their presence creates opportunities to study how additional exposures modify outcomes against a known molecular background. Although research into vascular mural-cell dysfunction and extracellular matrix abnormalities in CADASIL can inform hypotheses about sporadic SVD, the extent to which mechanisms and treatment responses generalize must be established empirically. The USCC’s combined clinical and biological assessments provide a setting in which to investigate these questions.

The harmonized protocol also provides a foundation for clinical trial preparation. High baseline completion rates for biospecimen collection and MRI demonstrate the feasibility of acquiring complementary measures across geographically dispersed centers. Follow-up at 18 and 36 months can establish their longitudinal variability and association with meaningful clinical outcomes. Those data will be needed to evaluate candidate prognostic biomarkers, determine suitable enrollment criteria, and estimate rates of change for future trials. Establishing a surrogate endpoint would require additional evidence that a treatment-induced change in the biomarker predicts clinical benefit. The present report documents the infrastructure and baseline population from which that work can proceed.

Several limitations warrant consideration. This cross-sectional analysis cannot establish progression or temporal relationships among manifestations. Recruitment through academic centers and affected families may preferentially include individuals with greater access to information and care. Predominantly White, highly educated participants and the exclusion of advanced disability constrain generalizability. Historical diagnoses and symptom dates are subject to recall error and incomplete documentation. Familial relatedness was unavailable for this analysis, and observations were treated as independent. Unmodeled clustering may affect standard errors and statistical inference. The smaller LR and MR groups limit between-tier comparisons. Finally, the number of outcomes examined increases the possibility of chance findings. Non-carrier family controls provide a valuable comparison group, but their shared backgrounds and experience of CADASIL distinguish them from population controls.

The USCC provides a harmonized national resource for studying variation in the clinical expression of pathogenic *NOTCH3* variants. Its baseline findings document cognitive and functional burden across risk tiers and establish the population for longitudinal investigation. Follow-up and coordinated comparisons with international cohorts will allow assessment of which genetic, biological, and environmental factors predict subsequent vascular injury and clinically meaningful change.

## Data Availability

All data produced in the present study are available upon reasonable request to the authors.

## Acknowledgments

This work was supported by the National Institutes of Health and Food and Drug Administration under award numbers RF1AG074608, R01AG082208, R01AG085602, and U01FD008399. This manuscript is the result of funding in whole or in part by the National Institutes of Health (NIH). The content is solely the responsibility of the authors and does not necessarily represent the official views of the National Institutes of Health. The authors acknowledge the research staff of the United States CADASIL Consortium: https://cadasil.wisc.edu/wp-content/uploads/sites/2133/2026/08/CADASIL_Consortium_Acknowledgments.pdf. The authors thank the valuable contributions of study participants and families to advance CADASIL research. The authors utilized the University of Wisconsin–Madison Biotechnology Center’s DNA Sequencing Facility (Research Resource Identifier – RRID:SCR_017759) to complete the described genomic analyses.

