## Supplemental Table 1 for "The United States CADASIL Consortium: Baseline Findings from a Natural History Study"

**Supplemental Table S1**

*Clinical Dementia Rating (CDR) Domain Ratings, Box Sum Scores, and Global Scores by CADASIL Pathogenicity Group*

|  | **CONTROL (N=116)** | **LOW (N=24)** | **MEDIUM (N=63)** | **HIGH (N=256)** | **Total (N=459)** | ***p*** |
| --- | --- | --- | --- | --- | --- | --- |
| **Memory**, n (%) |  |  |  |  |  | <.001^1^ |
| None | 87 (80.6%) | 12 (50.0%) | 33 (55.0%) | 113 (47.3%) | 245 (56.8%) |  |
| Questionable | 18 (16.7%) | 9 (37.5%) | 20 (33.3%) | 83 (34.7%) | 130 (30.2%) |  |
| Mild | 3 (2.8%) | 3 (12.5%) | 7 (11.7%) | 34 (14.2%) | 47 (10.9%) |  |
| Moderate | 0 (0.0%) | 0 (0.0%) | 0 (0.0%) | 7 (2.9%) | 7 (1.6%) |  |
| Severe | 0 (0.0%) | 0 (0.0%) | 0 (0.0%) | 2 (0.8%) | 2 (0.5%) |  |
| Missing | 8 | 0 | 3 | 17 | 28 |  |
| **Orientation**, n (%) |  |  |  |  |  | <.001^1^ |
| None | 99 (91.7%) | 18 (75.0%) | 46 (76.7%) | 172 (72.0%) | 335 (77.7%) |  |
| Questionable | 8 (7.4%) | 5 (20.8%) | 11 (18.3%) | 45 (18.8%) | 69 (16.0%) |  |
| Mild | 1 (0.9%) | 1 (4.2%) | 3 (5.0%) | 18 (7.5%) | 23 (5.3%) |  |
| Moderate | 0 (0.0%) | 0 (0.0%) | 0 (0.0%) | 3 (1.3%) | 3 (0.7%) |  |
| Severe | 0 (0.0%) | 0 (0.0%) | 0 (0.0%) | 1 (0.4%) | 1 (0.2%) |  |
| Missing | 8 | 0 | 3 | 17 | 28 |  |
| **Judgment and problem-solving**, n (%) |  |  |  |  |  | <.001^1^ |
| None | 100 (92.6%) | 17 (70.8%) | 43 (71.7%) | 149 (62.3%) | 309 (71.7%) |  |
| Questionable | 7 (6.5%) | 5 (20.8%) | 14 (23.3%) | 56 (23.4%) | 82 (19.0%) |  |
| Mild | 1 (0.9%) | 2 (8.3%) | 3 (5.0%) | 27 (11.3%) | 33 (7.7%) |  |
| Moderate | 0 (0.0%) | 0 (0.0%) | 0 (0.0%) | 6 (2.5%) | 6 (1.4%) |  |
| Severe | 0 (0.0%) | 0 (0.0%) | 0 (0.0%) | 1 (0.4%) | 1 (0.2%) |  |
| Missing | 8 | 0 | 3 | 17 | 28 |  |
| **Community affairs**, n (%) |  |  |  |  |  | <.001^1^ |
| None | 106 (98.1%) | 19 (79.2%) | 42 (70.0%) | 169 (70.7%) | 336 (78.0%) |  |
| Questionable | 2 (1.9%) | 4 (16.7%) | 17 (28.3%) | 51 (21.3%) | 74 (17.2%) |  |
| Mild | 0 (0.0%) | 1 (4.2%) | 0 (0.0%) | 12 (5.0%) | 13 (3.0%) |  |
| Moderate | 0 (0.0%) | 0 (0.0%) | 1 (1.7%) | 5 (2.1%) | 6 (1.4%) |  |
| Severe | 0 (0.0%) | 0 (0.0%) | 0 (0.0%) | 2 (0.8%) | 2 (0.5%) |  |
| Missing | 8 | 0 | 3 | 17 | 28 |  |
| **Home and hobbies**, n (%) |  |  |  |  |  | <.001^1^ |
| None | 103 (95.4%) | 19 (79.2%) | 41 (68.3%) | 152 (63.6%) | 315 (73.1%) |  |
| Questionable | 4 (3.7%) | 3 (12.5%) | 13 (21.7%) | 55 (23.0%) | 75 (17.4%) |  |
| Mild | 0 (0.0%) | 2 (8.3%) | 6 (10.0%) | 22 (9.2%) | 30 (7.0%) |  |
| Moderate | 1 (0.9%) | 0 (0.0%) | 0 (0.0%) | 8 (3.3%) | 9 (2.1%) |  |
| Severe | 0 (0.0%) | 0 (0.0%) | 0 (0.0%) | 2 (0.8%) | 2 (0.5%) |  |
| Missing | 8 | 0 | 3 | 17 | 28 |  |
| **Personal care**, n (%) |  |  |  |  |  | .044^1^ |
| None | 107 (99.1%) | 24 (100.0%) | 56 (93.3%) | 221 (92.5%) | 408 (94.7%) |  |
| Questionable | 1 (0.9%) | 0 (0.0%) | 0 (0.0%) | 2 (0.8%) | 3 (0.7%) |  |
| Mild | 0 (0.0%) | 0 (0.0%) | 4 (6.7%) | 12 (5.0%) | 16 (3.7%) |  |
| Severe | 0 (0.0%) | 0 (0.0%) | 0 (0.0%) | 4 (1.7%) | 4 (0.9%) |  |
| Missing | 8 | 0 | 3 | 17 | 28 |  |
| **Language**, n (%) |  |  |  |  |  | <.001^1^ |
| None | 100 (92.6%) | 19 (79.2%) | 46 (76.7%) | 157 (66.0%) | 322 (74.9%) |  |
| Questionable | 5 (4.6%) | 5 (20.8%) | 10 (16.7%) | 56 (23.5%) | 76 (17.7%) |  |
| Mild | 2 (1.9%) | 0 (0.0%) | 4 (6.7%) | 21 (8.8%) | 27 (6.3%) |  |
| Moderate | 1 (0.9%) | 0 (0.0%) | 0 (0.0%) | 3 (1.3%) | 4 (0.9%) |  |
| Severe | 0 (0.0%) | 0 (0.0%) | 0 (0.0%) | 1 (0.4%) | 1 (0.2%) |  |
| Missing | 8 | 0 | 3 | 18 | 29 |  |
| **Behavior**, n (%) |  |  |  |  |  | <.001^1^ |
| None | 100 (92.6%) | 22 (91.7%) | 51 (85.0%) | 176 (73.9%) | 349 (81.2%) |  |
| Questionable | 4 (3.7%) | 1 (4.2%) | 5 (8.3%) | 43 (18.1%) | 53 (12.3%) |  |
| Mild | 4 (3.7%) | 1 (4.2%) | 4 (6.7%) | 18 (7.6%) | 27 (6.3%) |  |
| Severe | 0 (0.0%) | 0 (0.0%) | 0 (0.0%) | 1 (0.4%) | 1 (0.2%) |  |
| Missing | 8 | 0 | 3 | 18 | 29 |  |
| **Motor**, n (%) |  |  |  |  |  | <.001^1^ |
| None | 104 (96.3%) | 21 (87.5%) | 43 (71.7%) | 171 (71.8%) | 339 (78.8%) |  |
| Questionable | 3 (2.8%) | 3 (12.5%) | 15 (25.0%) | 53 (22.3%) | 74 (17.2%) |  |
| Mild | 1 (0.9%) | 0 (0.0%) | 1 (1.7%) | 12 (5.0%) | 14 (3.3%) |  |
| Moderate | 0 (0.0%) | 0 (0.0%) | 1 (1.7%) | 1 (0.4%) | 2 (0.5%) |  |
| Severe | 0 (0.0%) | 0 (0.0%) | 0 (0.0%) | 1 (0.4%) | 1 (0.2%) |  |
| Missing | 8 | 0 | 3 | 18 | 29 |  |
| ^1^Kruskal-Wallis p-value | | | | | | |
